# Augmenting maternal clinical cohort data with administrative laboratory dataset linkages: a validation study

**DOI:** 10.1101/2024.06.19.24309149

**Authors:** Laura Rossouw, Nkosinathi Ngcobo, Kate Clouse, Cornelius Nattey, Karl-Günter Technau, Mhairi Maskew

## Abstract

**Background:** The use of big data and large language models in healthcare can play a key role in improving patient treatment and healthcare management, especially when applied to large-scale administrative data. A major challenge to achieving this is ensuring that patient confidentiality and personal information is protected. One way to overcome this is by augmenting clinical data with administrative laboratory dataset linkages in order to avoid the use of demographic information.

**Methods:** We explored an alternative method to examine patient files from a large administrative dataset in South Africa (the National Health Laboratory Services, or NHLS), by linking external data to the NHLS database using specimen barcodes associated with laboratory tests. This offers us with a deterministic way of performing data linkages without accessing demographic information. In this paper, we quantify the performance metrics of this approach.

**Results:** The linkage of the large NHLS data to external hospital data using specimen barcodes achieved a 95% success. Out of the 1200 records in the validation sample, 87% were exact matches and 9% were matches with typographic correction. The remaining 5% were either complete mismatches or were due to duplicates in the administrative data.

**Conclusions:** The high success rate indicates the reliability of using barcodes for linking data without demographic identifiers. Specimen barcodes are an effective tool for deterministic linking in health data, and may provide a method of creating large, linked data sets without compromising patient confidentiality.

## Background

The introduction of big data and large language models in health and healthcare research have the potential to contribute substantially to patient treatment and healthcare management. Linkages between large administrative datasets can improve clinical management by facilitating diagnosis and treatment across facilities. On the research side more advanced clinical modelling and detailed analyses of disease patterns and outbreaks have been enabled, creating opportunities to leverage routinely collected data into applicative information. ^1^

One major challenge facing the utilisation of large-scale administrative health data sources is the potential implication for patient confidentiality and protection of personal information. While de-identification of data, in addition to robust data security protocols (encryption, key-coding, virtual private networks etc.), have been standard data security measures for some time, national legislation to protect the personal information of populations represented in data is becoming more widespread practice. This includes the General Data Protection Regulation in Europe^2^, and the Protection of Personal Information (POPI) Act in South Africa.^3^ Here, we outline some challenges presented by data processing regulations in terms of the usability of big data and explore alternative methods to overcome this barrier without compromising patient confidentiality.

### Case study: The use of big data in HIV care management and research in South Africa

One example of how big data could be utilised effectively in a resource-constrained environment is in HIV care management. Although South Africa has made great strides in reducing new cases of HIV in the last 20 years, the number of people living with HIV in South Africa remains high by global standards. Findings from the 2022 National HIV Prevalence, Incidence, Behavior and Communication Survey (SABSSM) showed that HIV prevalence across all ages was 12.7%, or an estimated 7.8 million people.^4^ HIV prevalence rates are often higher among specific population subsets, including pregnant women, among whom the HIV prevalence rate is approximately 27.5%^5^. Several significant milestones in HIV management and care have been achieved since the roll-out of the national antiretroviral treatment (ART) programme in 2004.^6^ Specifically, ART coverage among pregnant women accessing antenatal care is near universal (>95%),^7^ and vertical transmission of HIV from mother to child at birth was as low as 0.7% in 2019.^8^

Despite these wins, the risk of transmission increases in the postnatal period, with the cumulative vertical transmission rate of 4.3% at 18 months of age. The increase in postnatal transmission rates peaks during the first six months of breastfeeding.^8^ This is a period which is particularly challenging for women and often results in poor adherence and retention in care, which negatively impacts viral suppression.^9,10^ As a result, this period has been identified as a priority in the prevention of vertical transmission of HIV.

In addition, there is a high rate of mobility among care recipients, with women often accessing care at different public healthcare facilities.^11^ Tracking care recipients during the antenatal and postnatal period is challenging due to lack of a national longitudinal HIV care database.^12^ Without a universal HIV care database, women moving between facilities are often considered lost to follow up, making assessment of the effectiveness of ART programmes problematic. It might also hamper care, as real-time data sharing about a care recipient between clinicians and providers at different facilities becomes near impossible.

### The challenge: using big data easily while protecting the privacy of care recipients

Creating a nationally representative cohort with unique identifiers is an important step towards effective HIV care management and program monitoring in South Africa. One attempt to create such a cohort for people living with HIV in South Africa involves using routinely collected laboratory data from the National Health Laboratory Services. The National Health Laboratory Services (NHLS) is the sole provider of laboratory and pathology services to the South African public sector, providing services to 80% of the South African population and data are stored at the NHLS Corporate Data Warehouse (CDW). Bor and colleagues applied probabilistic linkage approaches to the NHLS data to group tests under unique identifiers, creating a nationally representative cohort which can be used to indicate access to care, monitor continuity of care and describe treatment outcomes. ^13^ More recently, our team has aimed to replicate this approach among pregnant women living with HIV in South Africa^.14^

While probabilistic linkage approaches frequently require access to demographic data like patient name, surname and birth dates to be effectively implemented, these identifying data are not readily available to those wishing to utilise or link external datasets to these data resources. Drawing any information from these data sets or linking it to more descriptive hospital-level or study data means that care recipients need an identifier which can be used to link them without compromising their rights outlined in the POPI Act.

## Methods

### Addressing the challenge: using specimen barcodes as a deterministic tool to link patient episodes

We explored an alternative method to examine patient files from the NHLS and to link external data to the NHLS database, through the use of specimen barcodes associated with laboratory tests. These alphanumeric barcodes are allocated centrally by the NHLS, and affixed to blood test tubes. The barcode is allocated to the blood test tube, regardless of the number of tests performed on the tube. As such, one barcode might be associated with different test types but is unique to the individual person accessing care.

While specimen barcodes offer a deterministic way of performing data linkages without accessing demographic information, and could in theory have quite high linkage accuracy, the validity of this approach has not been quantified previously. Here we describe the validation of linkages between maternal cohort data and the NHLS National HIV cohort using laboratory barcode data and quantify the performance metrics of this approach. We also outline potential limitations and next steps to improve the usefulness of this technique.

### Description of the Data Used

This validation study utilised two data sources. First, we accessed laboratory data available in the NHLS National HIV Cohort. This longitudinal cohort comprises laboratory data from the NHLS. In 2018, this included approximately 94 million observations on 13 million individuals. The dataset includes information on the care recipient’s demographic information (name, surname, sex and date of birth), geography and facility visited, HIV monitoring tests (including CD4 count, viral load, and other test results).^14^

The second dataset used for validation emerges from routinely collected maternal data from the Rahima Moosa Maternal and Child Hospital (RMMCH) in Gauteng Province. RMMCH is a maternal hospital and academic paediatric referral center in Johannesburg, South Africa, offering both in-hospital admissions and outpatient facilities. In addition to maternal care history, the RMMCH cohort contains barcodes for all blood tests and specimens performed for the sample of care recipients, which in this analysis were pregnant women that visited RMMCH between 2013 and 2018.

### The Validation Process

Specimen barcodes were used to link the RMMCH data to the NHLS data for pregnant women in a deterministic fashion. To validate the accuracy of these linked records, a random subset (approximately 10%) of individuals were drawn from the matched sample. For this random subset, the researchers received permission from the Rahima Moosa Mother and Child Hospital and the Empilweni Services and Research Unit and the NHLS Office of Academic Affairs and Research to access demographic identifiers from each dataset, and were able to manually validate the success of matching on barcodes. This sample is referred to as the validation sample.

The validation sample was independently assessed by two researchers. Observations were broadly classified as (1) an exact match, (2) a match after correction of typographic errors, and (3) not a match. Observations in the exact match category are observations where the name, surname, date of birth and gender reported in both the RMMCH data and NHLS matched completely. The second category, matched with correction of typographic errors, usually had one or two errors, but the researcher could discern that it was in fact the same person. For category (3), the information on the mother was completely different from the RMMCH and NHLS. After the validation, two additional categories were created, namely (4) observations that were complete duplicates of other observations in the validation sample and (5) observations that did not contain enough demographic information to be placed in any of the earlier categories.

### Ethics

Analyses of data from the Rahima Moosa Mother and Child Hospital Maternal HIV Cohort and linkage of this cohort to laboratory data from the National Health Laboratory Services approved under protocol M200237 of the Human Research Ethics Committee (Medical) of the University of the Witwatersrand.

#### Validation results

1200 records were sampled and manually reviewed in the validation sample. The validation sample consists of the name, surname, date of birth and gender of the care recipients as it was reported in both the NHLS and the RMMCH sample. In linking patient barcodes in the NHLS to the RMMCH data, there was a linking success rate of 95% (n=1148) (Table 1).

**Table 1:**
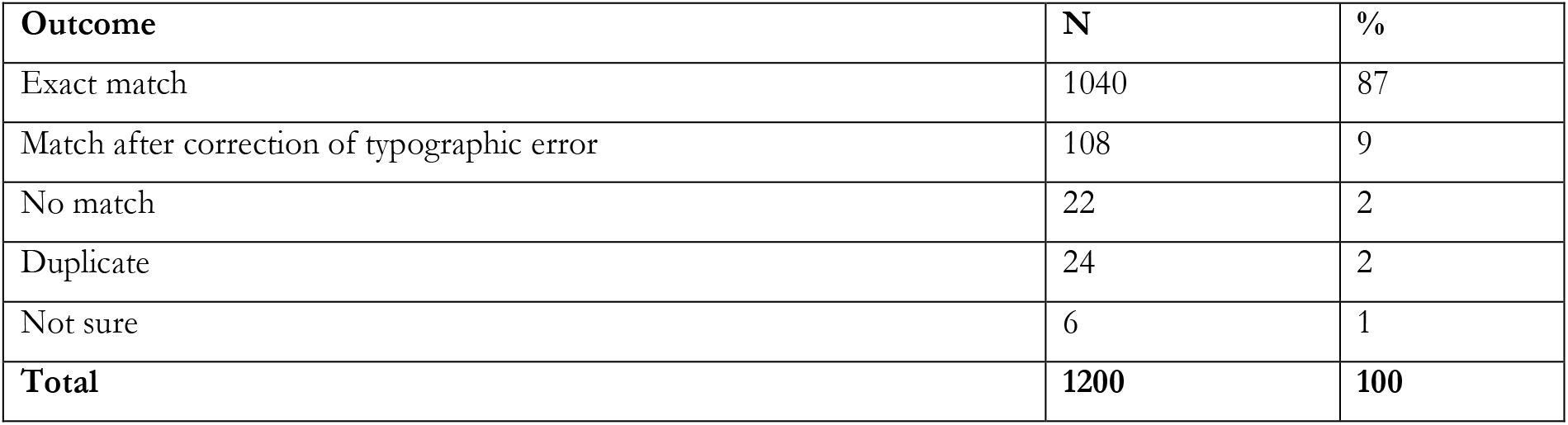
Results from barcode validation.

| Outcome | N | % |
| --- | --- | --- |
| Exact match | 1040 | 87 |
| Match after correction of typographic error | 108 | 9 |
| No match | 22 | 2 |
| Duplicate | 24 | 2 |
| Not sure | 6 | 1 |
| **Total** | **1200** | **100** |

- Most (n=1040) of these data points were exact matches, while 108 could be validated as matches, but had typographical errors when it came to the names, surnames, date of birth or gender. These are described in detail in Table 2. The majority of errors were in the form of date of birth (85%), while 22% of cases had errors in the name or surname, and 11% were errors based on gender. A matched observation could have more than one of these errors.
- Two percent (n=22) of the records were not matched and there was evidence that there were different people.
- Another 24 (2%) were duplicate observations, i.e. they had exact duplicates in the validation sample. These were instances where there were multiple observations for an individual in the NHLS cohort that matched with an individual in the RMMCH data.
- In six cases (1%), it was difficult to assess the validity of the match as information such as date of birth were missing and names clearly did not match. It is possible that some of these cases were instances where a mother’s name was attached to the barcode in one data file, while the infant’s name was linked to the barcode in the other data file.

## Discussion: Opportunities using this approach

The high percentage of successful linkages between the RMMCH data and the NHLS data using testing barcodes points to the reliability of using barcodes as a linking measure when creating a universal data set in South Africa. Creating this data set is key to tracking the pregnancy journey of women living with HIV, especially their movement across facilities. Another benefit of using the NHLS to map this journey is that the data are collected from the source testing platforms, which should in theory result in a smaller probability of data capturing error than other data collection techniques. In addition, data collection costs are kept low as data are collected from routine patient care rather than collecting national surveys.^15^ The data set can be used to investigate various relevant outcomes, including the effect of system-wide factors impacting the continuity of care among pregnant women, and the individual-level risk factors for loss from HIV care.^14^

**Table 2:** Type of typographic errors for the “match after correction of typographic error” category.

| Outcome: Match after correction of typographic error | N | % |
| --- | --- | --- |
| Error in the date of birth | 92 | 85 |
| Error in the name or surname | 24 | 22 |
| Error in the gender allocation | 11 | 10 |

## Data Availability

The external data underlying this article were provided with permission by the data gatekeeper for Rahima Moosa Mother and Child Hospital and the Empilweni Services and Research Unit. Cohort participants provided written consent for data to be used for research purposes and requests for access can be directed to Empliweni Services and Research Unit, Johannesburg. Laboratory data linked are owned by the National Health Laboratory Services and access is governed by policies and procedures in response to requests made directly to the NHLS Office of Academic Affairs and Research.

## Acknowledgements

We wish to acknowledge the research study teams at Rahima Moosa Mother and Child Hospital and the Empilweni Services and Research Unit who collected data on all cohort participants and provide on-going data quality support and access. We also wish to acknowledge the South African National Health Laboratory Services for access to laboratory data.

